# Residual tiny pulmonary vein potentials on high-resolution map after pulsed field ablation predicts late-phase reconnection between the left atrium and pulmonary vein

**DOI:** 10.1101/2025.08.28.25334689

**Authors:** Masaharu Masuda, Takashi Sumigawa, Hiroyuki Uematsu, Shota Kato, Hirotaka Ooka, Satoshi Kudo, Mizuki Ochi, Shin Okamoto, Takayuki Ishihara, Kiyonori Nanto, Takuya Tsujimura, Yosuke Hata, Sho Nakao, Masaya Kusuda, Wataru Ariyasu, Haruna Miyaguchi, Kohei Nanri, Toshiaki Mano

**Affiliations:** Kansai Rosai Hospital Cardiovascular Center, 3-1-69 Inabaso, Amagasaki, Hyogo 660-8511, Japan

**Keywords:** Atrial fibrillation, pulse field ablation, residual pulmonary vein potential, pulmonary vein isolation, endpoint

## Abstract

**Background:** Pulsed-field ablation (PFA) is becoming more widely used as its efficacy and safety. However, the durability of pulmonary vein isolation (PVI) is not necessarily satisfactory even with PFA. This study aimed to investigate whether post-PFA residual tiny pulmonary vein potentials (PVPs) on a high-resolution map may serve as a predictor of late-phase reconnection between the left atrium and pulmonary vein.

**Methods:** Fifteen patients who underwent PFA-based PVI, and the second ablation for recurrent atrial tachyarrhythmias were enrolled. The association between residual tiny PVPs (≥ 0.03 mV) on a post-PFA high-resolution map and reconnected PVPs observed during the second ablation were studied. The presence or absence of PVPs in 270 pulmonary vein segments from 15 cases was compared between the post-PFA map of the initial ablation and the map from the second ablation.

**Results:** Among 42 post-PFA residual tiny PVPs, 34 (81%) showed locational concordance with the late-phase reconnected PVPs identified at the second ablation. Thirty-four (52%) out of 66 late-phase reconnected PVPs at the second ablation were located at sites concordant with the post-PFA residual tiny PVPs. Post-PFA residual tiny PVPs defined as bipolar voltage of ≥ 0.03 mV well predicted late-phase reconnected PVPs (sensitivity=52%, specificity=96%, positive predictive value=81%, negative predictive value=86%).

**Conclusion:** The presence of post-PFA residual tiny PVPs can be used as late-phase reconnection between the left atrium and pulmonary vein.

**Condensed abstract:** This observational study aimed to investigate whether post-PFA residual tiny pulmonary vein potentials (PVPs) on a high-resolution map may serve as a predictor of late-phase reconnection between the left atrium and pulmonary vein. Fifteen patients were who underwent pulmonary vein isolation using pulse field ablation and the second ablation for recurrent atrial tachyarrhythmias were enrolled. Residual tiny PVPs defined as bipolar voltage of ≥ 0.03 mV well predicted late-phase reconnected PVPs (sensitivity=52%, specificity=96%, positive predictive value=81%, negative predictive value=86%). In conclusion, the presence of post-PFA residual tiny PVPs can be used as late-phase reconnection between the left atrium and pulmonary vein

## Introduction

Pulmonary vein isolation (PVI) is a cornerstone of atrial fibrillation (AF) ablation.^1^ Pulsed-field ablation (PFA) is becoming more widely used due to its established efficacy and safety.^2–4^ However, the durability of PVI is not necessarily satisfactory even with PFA.^5,6^ One of the reasons for this is that acute elimination of local electrogram signal by PFA does not coincide with the formation of durable lesions extending into the late phase.^5,7^ Therefore, in clinical practice, it is expected to elucidate the appropriate intraoperative endpoints for PFA to ensure durable PVI.

On a high-resolution voltage map after PVI with PFA, residual tiny electrogram signals are sometimes recorded within the pulmonary vein and its antrum, even without significant electrogram recordings by ablation catheter.^8^ Previous studies on radiofrequency-based PVI have demonstrated that targeting residual potentials observed in the antral region after extended PVI with additional ablation can improve long-term PVI durability.^9^ We hypothesize that the presence of post-PFA residual tiny pulmonary vein potentials (PVPs) observed on high-resolution mapping may suggest insufficient PFA in the surrounding areas, and may be associated with clinically-relevant reconnected PVPs in the late phase. The purpose of this study was to clarify the association between post-PFA residual tiny PVPs on a high-resolution map and reconnected PVPs at the second ablation in patients who underwent the second ablation for recurred atrial tachyarrhythmias.

## Methods

### Patients

From September 2024 to August 2025, consecutive patients who underwent a second ablation for recurrent AF, following PVI using either a penta-spline variable conformation PFA catheter (Boston Scientific, Marlborough [Cambridge] MA, USA) or the fixed-loop PulseSelect catheter (Medtronic, Mounds View, MN, USA), or the variable-loop Varipulse catheter (Biosense Webster, Diamond Bar, CA, USA) at the initial ablation. We excluded patients without high-resolution mapping on CARTO 3 (Biosense Webster) using an octa-spline catheter (Octaray, Biosense Webster) after PVI at the initial ablation and at the start of the second ablation. In addition, patients who underwent additional ablation after post-PFA high-resolution map was also excluded. Other exclusion criteria were inappropriate substrate mapping, age < 20 years, and prior cardiac surgery.

This study complied with the Declaration of Helsinki. The protocol was approved by our institutional review board, and written informed consent for the ablation and participation in the study was obtained from all patients.

### Ablation

Electrophysiological studies and catheter ablation were performed under general anesthesia using intravenous propofol at 5–10 ml as a bolus injection followed by 0.5 ml/kg/hr as a maintenance dose. A ventilator (HAMILTON C-1®; Hamilton Medical, Bonaduz, Switzerland) in synchronized intermittent mandatory ventilation mode with a respiratory rate of 12 breaths per min and tidal volume of 400 or 500 ml was used together with a supraglottic airway device (i-gel®; Intersurgical Limited, Berkshire, UK). A 6-Fr decapolar electrode catheter was introduced into the coronary sinus. Following transseptal puncture at the fossa ovalis, a deflectable long sheath specific to each pulsed field ablation (PFA) catheter—either the FARADRIVE (Boston Scientific) or the FlexCath (Medtronic)—was advanced into the left atrium.

PFA was performed using one of three single-shot PFA catheters: the pentaspline, catheter, the fixed-loop catheter, or the variable-loop catheter in accordance with the respective manufacturer’s instructions. The standard delivery protocol consisted of a minimum of eight PFA applications per pulmonary vein: for the FARAWAVE catheter, four applications in the “basket” configuration and four in the “flower” configuration; for the PulseSelect catheter, four applications at the pulmonary vein ostium and four at the antrum. When using a variable-loop catheter, a minimum of four applications were delivered per pulmonary vein: two at the ostium and two at the antrum. Additional applications beyond the standard eight were allowed at the discretion of the operator. Pulmonary vein and antral potentials were assessed using an ablation catheter, and additional PFA applications were delivered until no residual potentials were detectable with the ablation catheter.

### High-resolution mapping

High-resolution mapping was performed following PVI at the initial ablation and at the start of the second ablation. If AF persisted at the timing of the mapping, electrical cardioversion was applied to restore sinus rhythm. The mapping was performed under pacing from the high right atrium at a rate of 100 ppm using automatic point acquisition module CARTO CONFIDENCE^®^ and Smart index^®^ (BiosenseWebster). Fast anatomical mapping volume was set at 17 on a scale of 1–20, and respiration correction was applied. Operators manipulated the multielectrode catheter to cover PV antrum and ostium in addition to the whole left atrium at a density which ensured a fill threshold of 5 mm. Bandpass filtering was performed at 16–500 Hz for the bipolar signals. A window of interest was defined prior to the QRS complex to identify atrial activity.

### Analysis of voltage map

The obtained voltage map was analyzed offline by a specialized clinical engineer. Post-PFA residual tiny PVPs were defined as electrograms with amplitudes ≥ 0.03 mV, which was determined based on the bipolar noise level observed when using the penta-spline catheter in the electrophysiology laboratory. For evaluation, the lower limit of the voltage range on the voltage map was set to 0.03 mV, and areas within the pulmonary veins exhibiting voltages above this threshold were identified. To validate the appropriateness of the 0.03 mV threshold, the same assessment was concurrently performed using alternative criteria: a voltage cutoff of ≥ 0.10 mV or the presence of a positive area of complex signal indicator (CSI).

Furthermore, reconnected PVPs were defined as areas with apparent signal amplitude widely observed within the pulmonary veins during the second ablation procedure. These areas were carefully examined—using pacing and other techniques—to distinguish far-field potentials, and were ultimately deemed by the operator to represent clinically significant PVPs that were successfully eliminated through ablation.

The anatomical locations of the identified potentials were classified into four regions per pulmonary vein—roof, anterior, posterior, and bottom—as well as the carina, resulting in a total of nine regions per the ipsilateral superior and inferior veins (Figure 1). For each region, the presence or absence of potentials, the maximum voltage, and the area of electrogram recording were quantitatively assessed.

**Figure 1.**
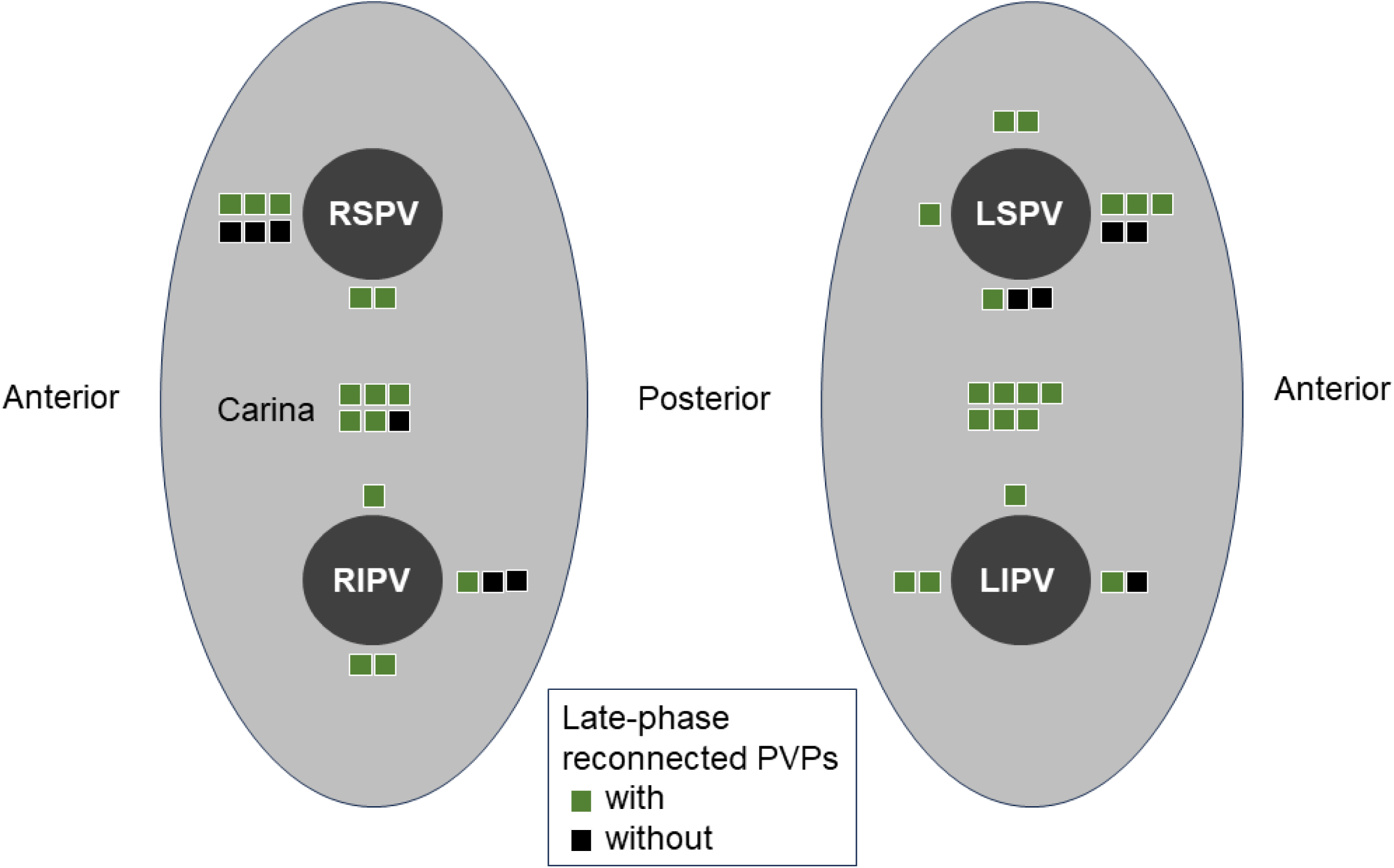
Post-PFA residual PVPs at the initial ablation. The distribution of residual PVPs recorded on high-resolution post-PFA map is shown. Each square indicates the presence of residual PVPs with bipolar voltage ≥ 0.030 mV. Cases with late-phase reconnected PVPs at the second ablation is highlighted by green color. PFA, pulse filed ablation; PVPs, pulmonary vein potentials.

### Follow-up

Patients were followed up every 2 to 4 weeks at the dedicated arrhythmia clinic of our institution. Routine ECGs were obtained at each outpatient visit, and 7-day ambulatory Holter monitoring was performed at 6 months post-ablation. When patients experienced symptoms suggestive of an arrhythmia, a surface ECG, ambulatory ECG, and/or cardiac event recording were also obtained. No antiarrhythmic drugs were prescribed 3 months post-ablation unless recurrent atrial tachyarrhythmias were observed. Patients with atrial arrhythmias recurrence were recommended to undergo the second ablation

### Statistical analysis

Continuous data are expressed as the mean ± standard deviation or median. Categorical data are presented as absolute values and percentages. Tests for significance were conducted using the unpaired t-test or nonparametric test (Mann-Whiney U test) for continuous variables, and the chi-square test or Fisher’s exact test for categorical variables. All analyses were performed using SPSS version 15.0 software (SPSS, Inc., Chicago, IL, USA).

## Results

### Patient and procedural characteristics

A total of 15 patients were included in the study. In all cases, the initial ablation procedure consisted solely of PVI using PFA catheter, and was completed without any complications. Among the arrhythmia recurrences, 10 cases were persistent AF, 2 were paroxysmal AF, and 3 were atrial tachycardia. A second ablation procedure was performed at a mean interval of 4.7 months (range: 2 to 7 months) following the initial ablation. Patients and procedural characteristics are shown in Table 1 and 2.

**Table 1.** Patient characteristics.

| Variable | n=15 |
| --- | --- |
| Age, years | 70 ± 9 |
| Female, n (%) |  |
| Body mass index, kg/m<sup>2</sup> | 25.3 ± 3.9 |
| Type of atrial fibrillation, n (%) |  |
| *Paroxysmal* | 5 (33) |
| *Persistent* | 10 (67) |
| Hypertension, n (%) | 7 (47) |
| Diabetes mellitus, n (%) | 3 (20) |
| History of heart failure, n (%) | 4 (27) |
| History of stroke/thromboembolism, n (%) | 3 (20) |
| Vascular disease, n (%) | 1 (7) |
| CHA<sub>2</sub>DS<sub>2</sub>-VASc | 2.4 ± 1.9 |
| NT-pro BNP, pg/ml | 767 (395, 1345) |
| Left atrial volume, ml | 94.9 ± 26.7 |
| Left superior pulmonary vein |  |
| *Maximum diameter, mm* | 23.7 ± 2.6 |
| *Minimum diameter, mm* | 19.8 ± 3.9 |
| Left inferior pulmonary vein |  |
| *Maximum diameter, mm* | 18.4 ± 3.1 |
| *Minimum diameter, mm* | 12.6 ± 3.6 |
| Right superior pulmonary vein |  |
| *Maximum diameter, mm* | 24.3 ± 3.1 |
| *Minimum diameter, mm* | 20.7 ± 3.5 |
| Right inferior pulmonary vein |  |
| *Maximum diameter, mm* | 20.2 ± 4.4 |
| *Minimum diameter, mm* | 13.5 ± 4.7 |

NT-pro BNP, N-terminal prohormone of brain natriuretic peptide

**Table 2.**
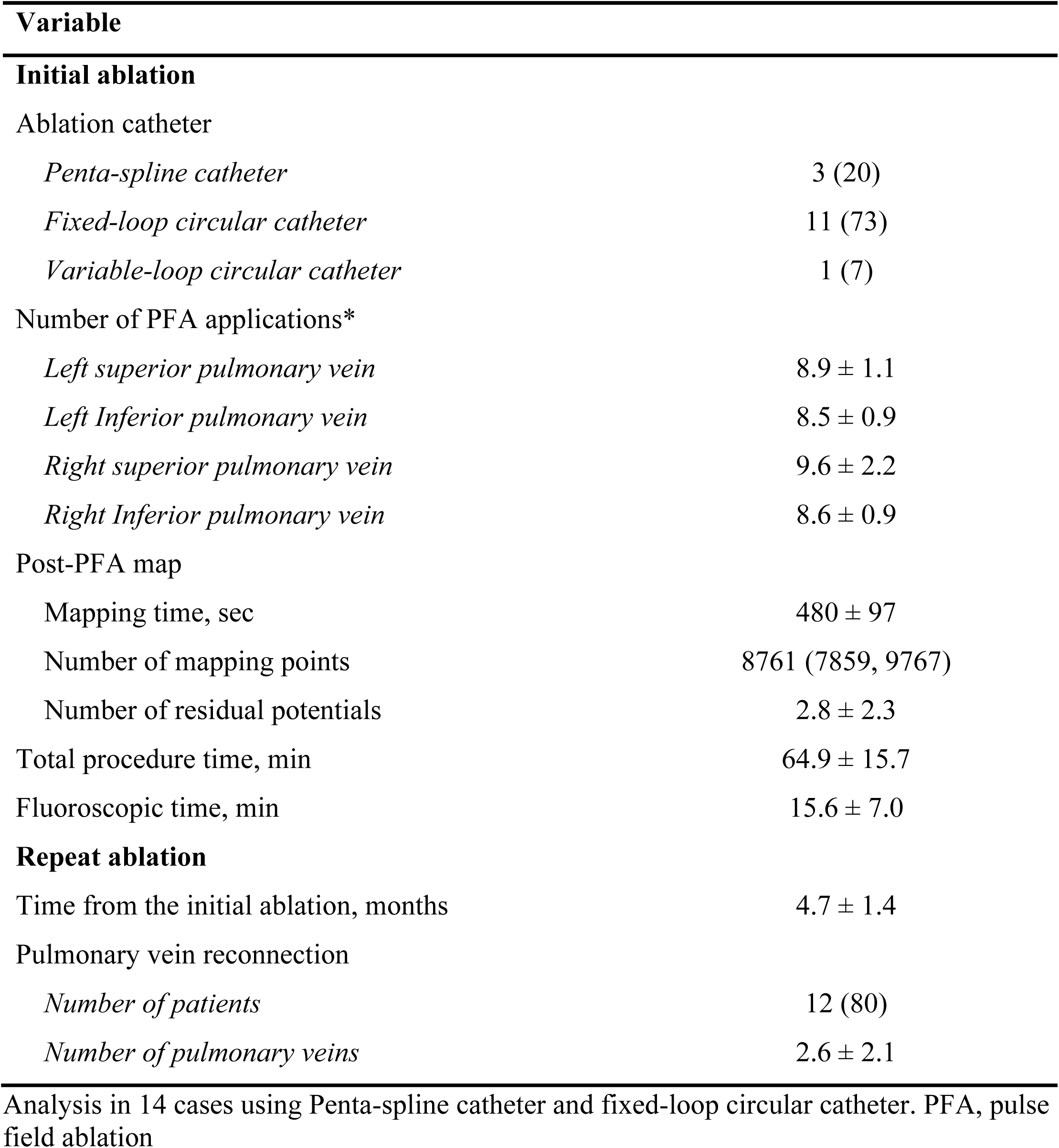
Ablation procedures.

| Variable |  |
| --- | --- |
| **Initial ablation** |  |
| Ablation catheter |  |
| *Penta-spline catheter* | 3 (20) |
| *Fixed-loop circular catheter* | 11 (73) |
| *Variable-loop circular catheter* | 1 (7) |
| Number of PFA applications* |  |
| *Left superior pulmonary vein* | 8.9 ± 1.1 |
| *Left Inferior pulmonary vein* | 8.5 ± 0.9 |
| *Right superior pulmonary vein* | 9.6 ± 2.2 |
| *Right Inferior pulmonary vein* | 8.6 ± 0.9 |
| Post-PFA map |  |
| Mapping time, sec | 480 ± 97 |
| Number of mapping points | 8761 (7859, 9767) |
| Number of residual potentials | 2.8 ± 2.3 |
| Total procedure time, min | 64.9 ± 15.7 |
| Fluoroscopic time, min | 15.6 ± 7.0 |
| **Repeat ablation** |  |
| Time from the initial ablation, months | 4.7 ± 1.4 |
| Pulmonary vein reconnection |  |
| *Number of patients* | 12 (80) |
| *Number of pulmonary veins* | 2.6 ± 2.1 |

Analysis in 14 cases using Penta-spline catheter and fixed-loop circular catheter. PFA, pulse field ablation

### Post-PFA residual tiny PVPs and late-phase reconnected PVPs

Post-PFA residual tiny PVPs at the initial ablation were predominantly distributed in the carina and anterior wall regions of both the left and right pulmonary veins (Figure 1). Among 42 post-PFA residual tiny PVPs, 34 (81%) showed locational concordance with the late-phase reconnected PVPs identified at the second ablation. Representative case demonstrating the locational concordance between post-PFA residual tiny PVPs and late-phase reconnected PVPs is shown in Figure 2. The distribution of late-phase reconnected PVPs is shown in the Figure 3. Thirty-four (52%) out of 66 reconnected PVPs were located at sites concordant with the post-PFA residual tiny PVPs.

**Figure 2.**
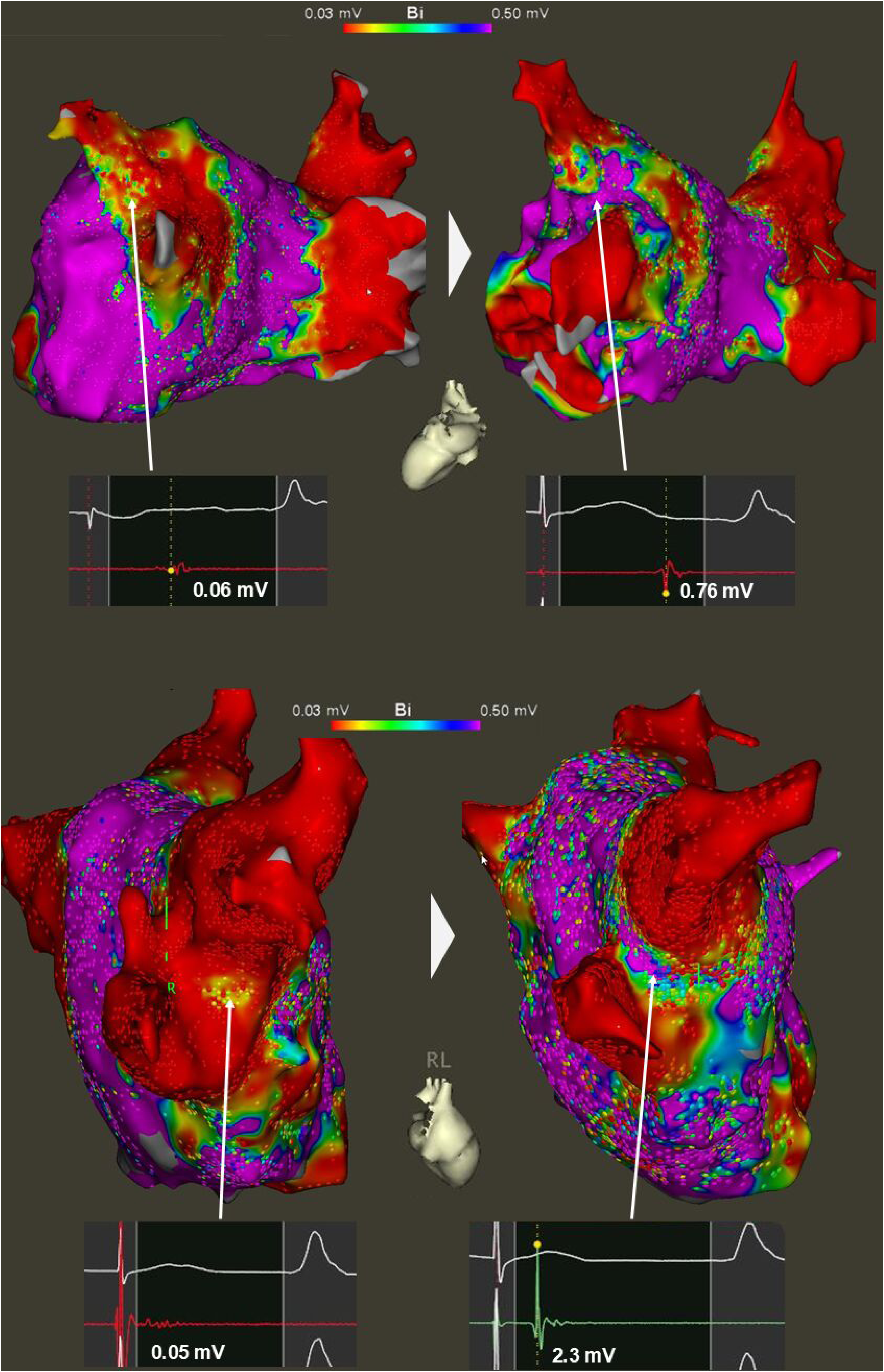
Representative cases showing concordance between post-PFA residual PVPs and late-phase reconnected PVPs. Upper case: A case of paroxysmal AF who underwent PVI using a fixed-loop circular PFA catheter. Residual PVPs were observed in the carinal region of left pulmonary vein. Five months later, during a second ablation procedure for recurrent AF, clearly reconnected and high-amplitude PVPs were identified in the same region. Lower case: A case of paroxysmal atrial fibrillation underwent PVI using a penta-spline catheter. Residual PVPs were observed on the anterior aspect of the right PV carina. Four months later, during a second ablation procedure for recurrent AF, high-amplitude and clearly reconnected PVPs were identified in the same region, with activation propagating from the anterior left atrial wall. AF, atrial fibrillation; PVI, pulmonary vein isolation; PFA, pulse filed ablation; PVPs, pulmonary vein potentials.

**Figure 3.**
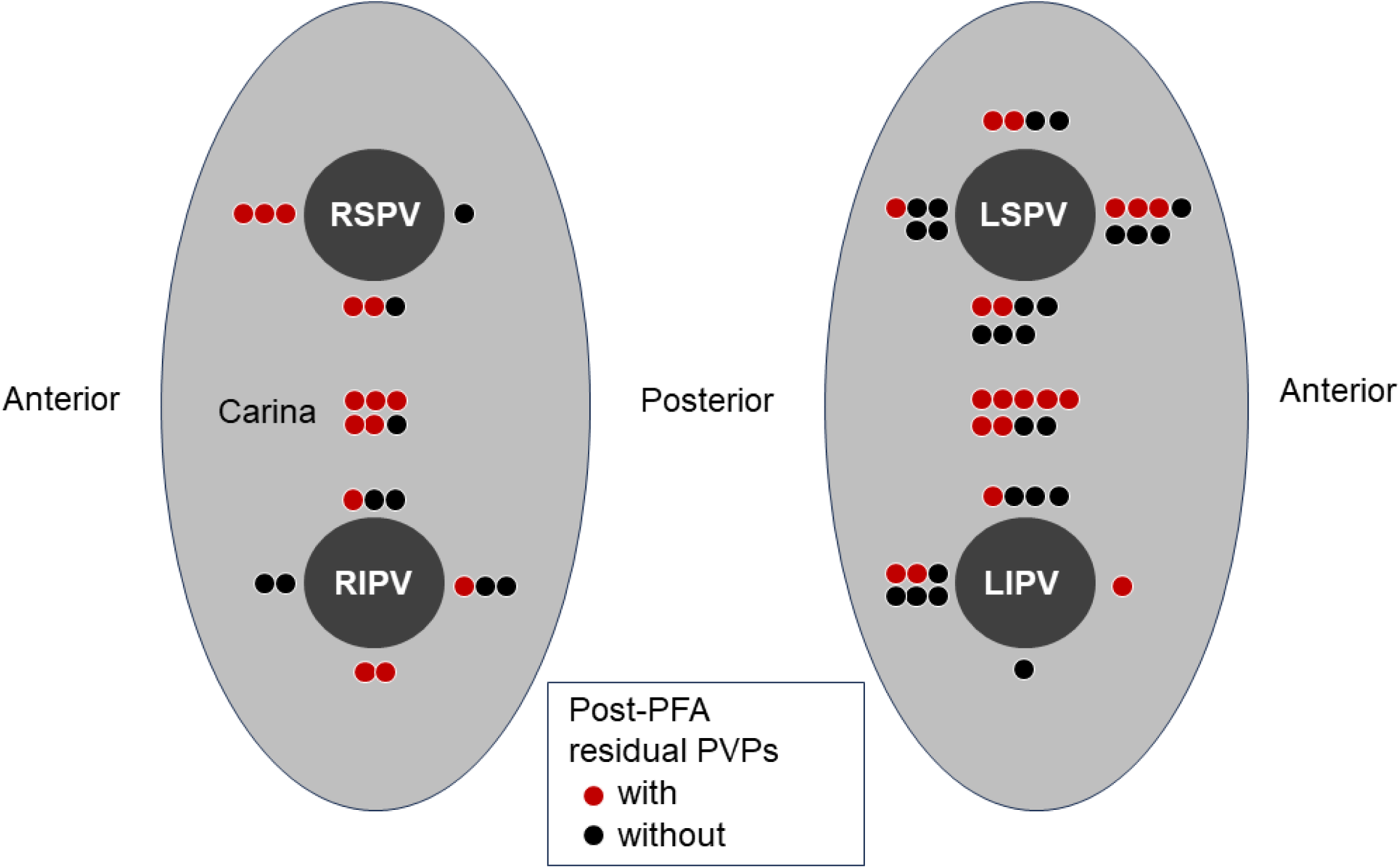
Late-phase reconnected PVPs on at the second ablation. The distributions of late-phase reconnected PVPs are shown by circles. Cases with post-PFA residual PVPs at the initial ablation are indicated by red color. PFA, pulse filed ablation; PVPs, pulmonary vein potentials.

### Signal characteristics of post-PFA residual PVPs

The areas where post-PFA residual tiny PVPs were recorded measured 0.73 ± 0.44 cm². The maximum bipolar voltage within each recording site was 0.30 ± 0.21 mV. There were no significant differences in either the recorded area size or the maximum bipolar voltage between post-PFA residual tiny PVPs that were concordant with late-phase reconnected PVPs and those that were not (Area, 0.78 ± 0.46 cm^2^ vs. 0.55 ± 0.33 cm^2^, p=0.198; Voltage, 0.31 ± 0.21 mV vs. 0.31 ± 0.18 mV, p= 0.969). Figure 4 illustrates the distribution of the area and maximum bipolar voltage of post-PFA residual tiny PVPs.

**Figure 4.**
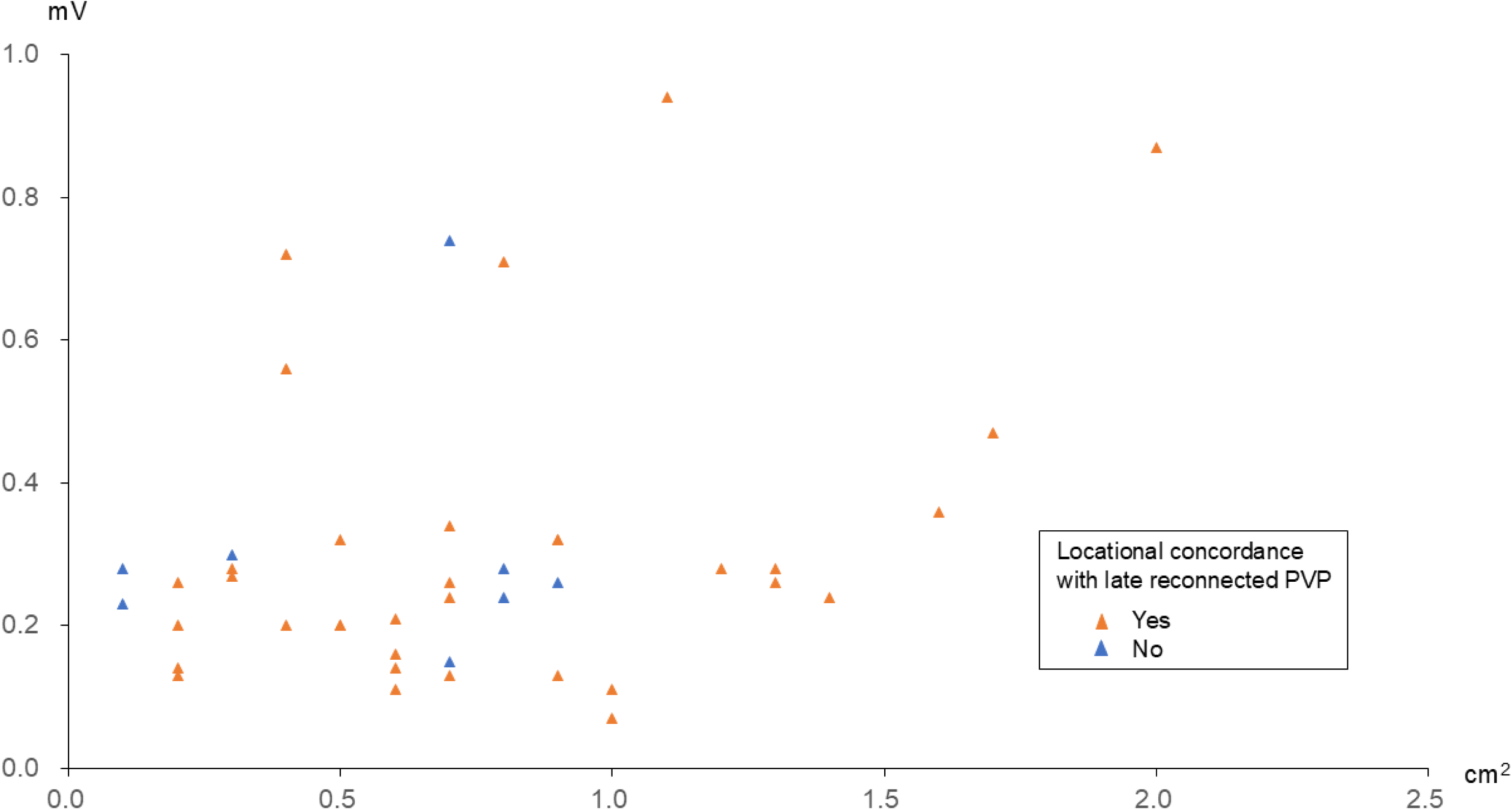
Signal characteristics of post-PFA residual PVPs. The distribution of area with post-PFA residual PVPs and the maximum bipolar voltage is indicated by triangles. The concordance between residual PVPs and late-phase reconnected PVPs is shown in orange. PFA, pulse filed ablation; PVPs, pulmonary vein potentials.

### Validity of the definition of residual PVPs

To validate the definition of post-PFA residual tiny PVPs from the perspective of its association with the presence of late-phase reconnected PVPs, the predictive accuracy of post-PFA residual tiny PVPs for late-phase reconnected PVPs was evaluated using data from 15 cases and 270 pulmonary vein segments (Figure 5). Three criteria were evaluated: bipolar voltage ≥ 0.03 mV, bipolar voltage ≥ 0.10 mV, and CSI positivity. Among these, a bipolar voltage threshold of ≥ 0.03 mV demonstrated the highest predictive accuracy for late-phase reconnected PVPs, with particularly high specificity.

**Figure 5.**
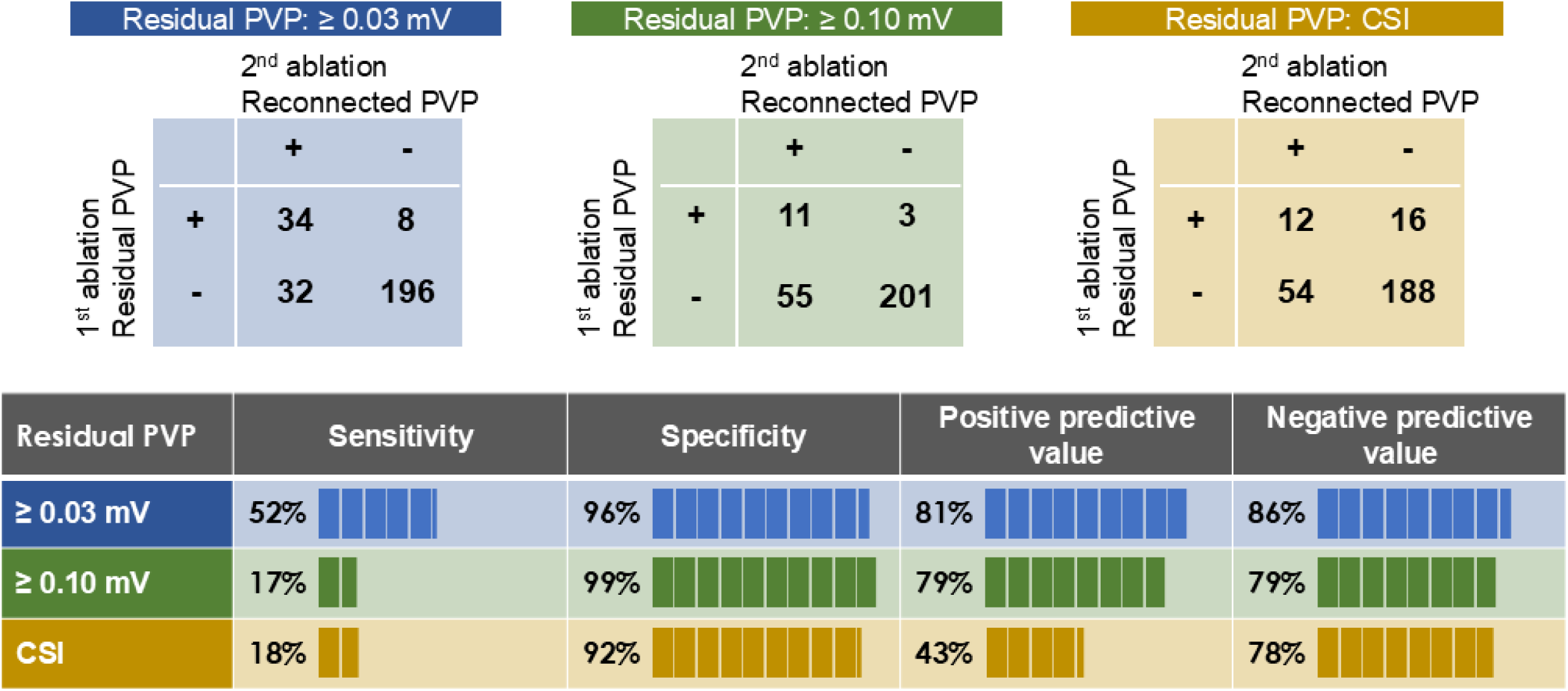
Predictive accuracy of various definitions of PVPs. The predictive accuracies of residual PVPs for the prediction of late-phase reconnected PVPs. The presence or absence of PVPs in 270 pulmonary vein segments from 15 cases was compared between the post-PFA map of the initial ablation and the map from the second ablation, in order to assess concordance or discordance. Three definitions of residual PVPs were employed: bipolar voltage ≥ 0.03 mV, bipolar voltage ≥ 0.10 mV, and CSI positive. PFA, pulse filed ablation; PVPs, pulmonary vein potentials; CSI, complex signal indicator.

## Discussion

This observational study investigated the association between post-PFA residual tiny PVPs on high-resolution map and late-phase LA-PV reconnections in patients who underwent the initial ablation using PFA ablation device and second ablation for recurrent atrial tachyarrhythmias. Major findings were as follows: (1) Despite the apparent elimination of electrical potentials as assessed by the PFA catheter, high-resolution mapping often identified residual, low-amplitude potentials persisting within the pulmonary veins and antral regions. (2) These residual potentials were more frequently observed in the carina regions and the anterior aspects of the superior PVs in both the left and right PVs. (3) The presence of post-PFA residual tiny potentials defined as a bipolar voltage of ≥ 0.03 mV, was correlated with late-phase LA-PV reconnection.

### Residual tiny PVPs on post-PFA high-resolution map as a predictor of poor long-term PVI durability

It is known that in PFA, there can be a discrepancy between the areas of acute electrogram elimination and those observed in the chronic phase.^7^ Therefore, the achievement of acute PVI does not necessarily guarantee durable PVI over the long term. A possible theoretical explanation is that, due to the unique characteristics of PFA, there tends to be a wider peripheral zone surrounding the area of irreversible tissue injury where transient electrogram suppression is observed.^10^ However, electrical activity in these regions may recover over time, more frequently than with conventional thermal ablation. Clinically, this phenomenon is thought to occur in areas surrounding the PFA lesion as well as in regions where the catheter had suboptimal contact with the myocardial tissue during ablation. As a result, PVI durability remains suboptimal although various strategies have been proposed to address and minimize these issues such as increasing the number of energy applications,^11,12^ confirming catheter– tissue contact using intracardiac echocardiography,^13^ and using impedance rise as an indicator of adequate contact.^14^

Considering the aforementioned context, establishing reliable acute-phase endpoints of PVI has become a critical issue for improving long-term PVI durability. In this study, residual tiny PVPs immediately after PVI were predictive of poor long-term PVI durability. Conversely, sites of late reconduction were frequently associated with the presence of tiny PVPs observed immediately after PVI. The presence of residual tiny PVPs suggests the existence of small regions of surviving electrical activity, indicating that ablation may not have achieved sufficient lesion intensity with continuous, transmural coverage. Given that residual electrical activity observed after PFA may expand over time and even involve adjacent healthy left atrial myocardium,^7,10^ it is plausible that residual potentials in the pulmonary vein antrum are associated with late left atrium-pulmonary vein reconnection.

### Technical consideration for the identification of tiny residual potentials

In this study, the CARTO system and the Octaray catheter were utilized to identify post-PFA tiny residual PVPs. The Octaray catheter features eight radiating flexible splines, making it well-suited for contact mapping of the pulmonary vein antrum, the carina, and within the pulmonary veins. Furthermore, its small electrode size (0.5 mm) and narrow interelectrode spacing (center-to-center distance of 3 mm) are considered optimal for the detection of low-amplitude signals. To identify residual tiny potentials predictive of late-phase LA-PV reconnection, several definitions of residual potentials were explored. Regarding the cutoff value for bipolar voltage, using the lowest feasible threshold close to the level of bipolar noise (0.03 mV) demonstrated superior sensitivity. The complex signal indication (CSI) module of the CARTO system, originally designed to identify slow conduction isthmuses in atrial tachycardia, demonstrated markedly inferior performance in both sensitivity and positive predictive value compared to the use of a bipolar voltage cutoff of 0.03 mV.

### Clinical implication

The presence of post-PFA residual tiny PVPs has been shown to predict late-phase reconnected PVPs. Assessment of residual tiny PVPs by high-resolution mapping following PFA-PVI, and elimination of recorded residual PVPs by additional PFA may serve as an acute endpoint for PVI, potentially contributing to the achievement of durable PVI. Whether additional ablation impacts PVI durability or AF recurrence remains an important subject for future investigation.

### Limitations

Several limitations of this study warrant mention. First, since this study included patients with AF recurrence and a second ablation procedure, there is substantial bias in the patient background. Therefore, caution should be exercised when generalizing these results to all patients undergoing initial AF ablation. Second, this is a small-scale, single-center study, and the results are likely to be significantly influenced by the operators’ proficiency in PFA ablation and mapping techniques.

## Conclusion

The presence of post-PFA residual tiny PVPs can be used as late-phase reconnection between the left atrium and pulmonary vein. The complete elimination of residual tiny PVPs may serve as a novel endpoint for PVI using PFA, further validation through randomized controlled trials is required.

## Data Availability

Data will be available on request to the correspoding author.

## Abbreviation list

PVI: pulmonary vein isolation
AF: atrial fibrillation
PFA: pulse field ablation
PVPs: pulmonary vein potentials

---

**Central illustration.**
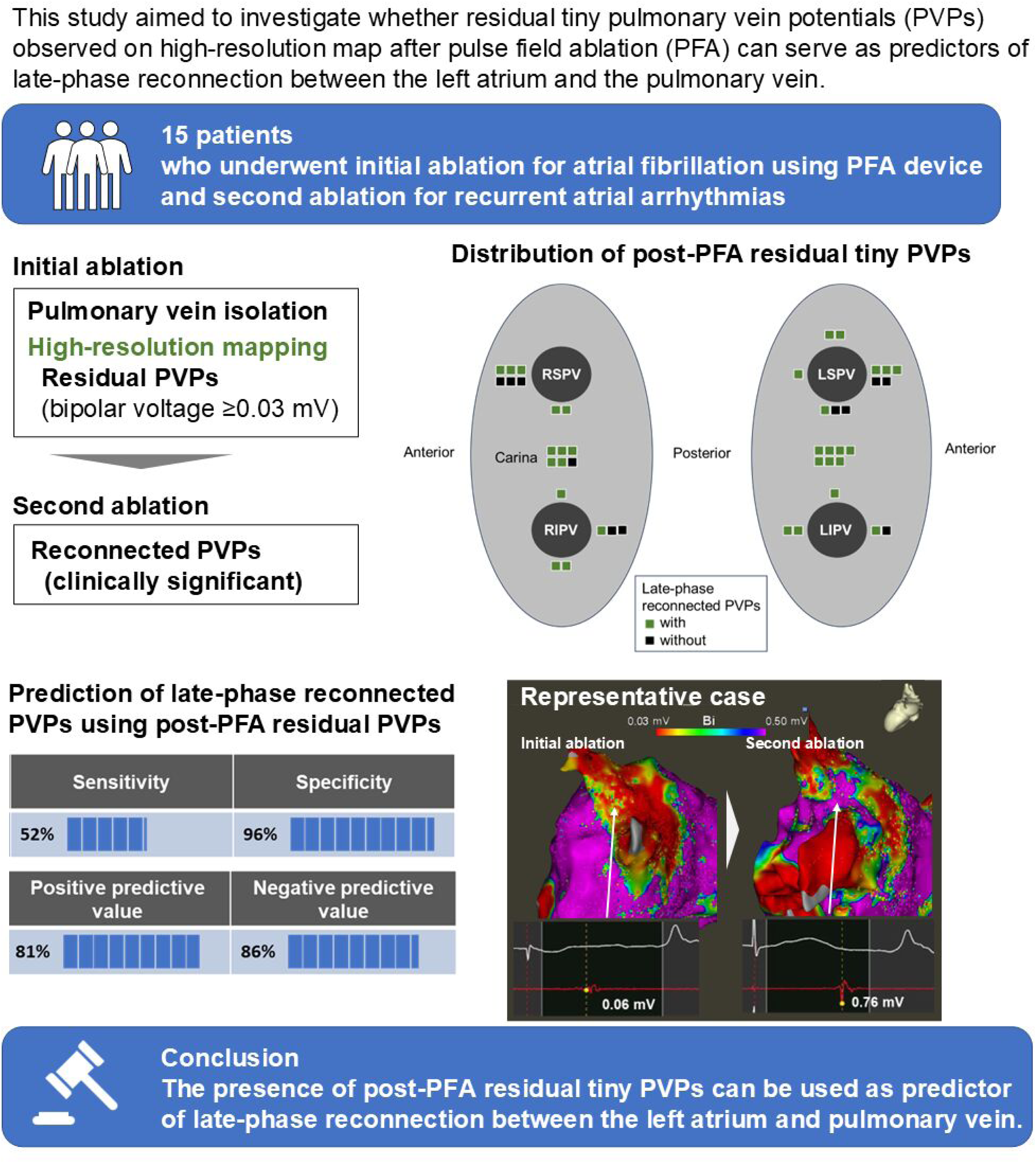
Abnormal LA substrates and accumulation of risk factors. Study design, representative maps demonstrating abnormal substrates, and AF recurrence rates stratified by the number of risk factors are shown. Accumulation of these risk factors increased AF recurrence rates in a stepwise manner with an HR = 1.90, 95% CI = 1.44 - 2.52, p<0.00001, for 1 risk increase. AF indicates atrial fibrillation; HR, hazard ratio; CI, confidence interval.

